# Assessment of absorbed dose in the hematopoietic stem cell layer of the bone marrow, assuming non-uniform distribution around the vascular endothelium of the bone marrow

**DOI:** 10.1101/2024.10.19.24315800

**Authors:** Noriko Kobayashi

## Abstract

Recent studies have shown that hematopoietic stem cells (HSCs) are concentrated around the endothelium of the sinusoidal capillaries. However, the current International Commission on Radiological Protection (ICRP) dosimetry model does not take into account the heterogeneity of the bone marrow tissue and stem cell distribution. In this study, the absorbed doses to the stem cell layer in the perivascular HSC layer in the bone marrow from radionuclides inhaled into the body were estimated for the major beta- and alpha-nuclides and noble gases and compared with the absorbed dose based on the ICRP 60 and ICRP 103 recommendations. The absorbed doses to the bone marrow obtained from the model calculations were not significantly different from ICRP60 and ICRP103 for beta-nuclides but were much lower than previously estimated for alpha-nuclides, and the contribution of red bone marrow and blood sources was greater than that of trabecular bone for alpha-nuclides. Noble gases in the red bone marrow may also affect the bone marrow stem cell layer. The bone marrow dose assessment for alpha nuclides and noble gases should be re-examined using a precise model based on CT images from the perspective of occupational and public radiation protection.

## INTRODUCTION

Bone marrow is one of the most radiosensitive organs; therefore, dose assessment assuming a bone microstructure and heterogeneous distribution of bone marrow tissues and cells is critical. The International Commission on Radiological Protection (ICRP) model currently adopted in Japan (ICRP publ.60)^(1)^ assumes a homogeneous distribution of trabecular bone tissues and bone marrow stem cells. Computational voxel phantoms have been introduced since the 2007 ICRP recommendation (ICRP 103)^(2)^, and a precise skeletal model developed by Hough et al.^(3)^ using micro-CT images of the trabecular spongiosa from an adult male cadaver has been incorporated into ICRP 133^(4)^, but hematopoietic stem cells (HSCs) are assumed to be uniformly distributed within the marrow cavities of hematopoietic active marrow^(5)^. The surrogate target for progenitor cells is specified as 10 μm from the bone surface in ICRP 60 and as “endosteum” 50 μm from the bone surface in ICRP 133, but the location of HSCs is not specified. However, recent studies have shown that HSCs and immune cells are localized around the endothelium of the bone marrow vessels^(6)^. One study reported that 85% of HSCs were located within 10μm of the bone marrow sinusoids in mice^(7)^. Kristensen et al.^(8)^ identified the microenvironment of HSCs and progenitors in the bone marrow using immunofluorescence staining of bone marrow tissue obtained from healthy volunteers. They found that the microenvironment of the HSCs was significantly enriched in sinusoids and megakaryocytes, whereas that of the progenitors was significantly enriched in capillaries, bone surfaces, and arteries. Therefore, it is necessary to re-evaluate the bone marrow dose, assuming that the HSC layer is localized around sinusoidal capillaries of the bone marrow. Especially for short-range alpha particles, the energy deposited in the target HSC layer can vary significantly depending on the distance from the source region.

Several bone marrow models have been developed for dosimetry of alpha-emitting radiopharmaceuticals, considering the microstructure of bone marrow tissue. Hobbs et al.^(9)^ developed a simple geometric model of marrow cavities considering the distribution of bone marrow cells. They calculated the absorbed doses from ^223^Ra in the trabecular bone surface or in the endosteal layer and found that the absorbed dose was predominantly deposited near the trabecular surface and ‘differed markedly from a standard absorbed fraction method.’ Tranel et al. ^(10)^ developed a cylindrical voxel bone marrow model with a blood vessel embedded in the center of the marrow and found that ‘the absorbed dose to the trabecular bone drops off quickly with increasing distance from the vessel wall because the range of alphas ensures that the absorbed dose is minimal at distances greater than 100μm.’ However, both studies assumed homogenous distribution of HSCs in the bone marrow cavity. Dosimetry, considering the arrangement of blood vessels in the bone marrow when the source is intravascular, remains a challenge.

In this study, a geometric model of trabecular bone and bone marrow tissue was constructed at the ㎛ scale, assuming that the HSC layer was localized in the perivascular HSC layer of the sinusoids. The absorbed doses of the stem cell layer from blood and trabecular bone sources were then estimated for selected beta nuclides, alpha nuclides, and noble gases and compared with the SAF values of ICRP 60 and 103. This is the first attempt at bone marrow dosimetry, assuming that the HSC layer is located around the sinusoidal capillaries in the bone marrow.

## 1. METHODS

### 2.1 Geometric modelling of trabecular bone and bone marrow tissues

A model of the trabecular bone tissues in the cervical vertebrae was created based on the data from JM-103 in the Japan Atomic Energy Agency (JAEA) Data/Code 2014-017^(11)^, using the PHITS code ver. 3.17^(12)^. The JM-103 data were used because a detailed weight breakdown of the bone tissue and blood was not available in the ICRP89^(13)^.

The height of the cervical vertebrae was estimated to be 9 cm based on the following assumptions: height 171 cm, length of the spine 52 cm (approximately 3/10 of the height), and cervical, thoracic, and lumbar vertebrae ratio of approximately 2:7:3. The weight of the bone tissue and blood in the cervical spine was calculated by summing the values given in Tables C1–C5 of the JAEA Data/Code 2014-017^(11)^. As the percentage of blood contained in each bone tissue was not reported, the amount of blood contained in the red bone marrow was calculated as 13.5% of the red bone marrow based on the percentages of the data reported in ICRP 89^(12)^ (7% of total blood distributed in bone tissue and 4% for blood distributed in the red bone marrow) (Table 1). Data on the percentage of blood distributed in the sinusoids of the blood distributed in the red bone marrow were not available; therefore, this was calculated to be 89.4%, as shown in Table 2, using data from mouse bone marrow vessels by Bixel et al. ^(14)^. The material densities were set at 1.765 g/cm^3^ for the trabecular bone ^(9)^ and 1 g/cm^3^ for the red bone marrow (RBM), soft tissues, and blood.

**Table 1.** Weight of JM-103 cervical bone tissues.

| Organ ID and name | Total body tissue (g)<br>※1 | Cortical bone(g) | Trabecular bone (g) | Soft tissues (g) | RBM (g)<br>※2 | Blood (g) | Blood in RBM (g)<br>※3 |
| --- | --- | --- | --- | --- | --- | --- | --- |
| 140 Cervical vertebra_01 | 0.8 |  | 0.2 | 0.6 | 0.5 | 0.1 | 0.1 |
| 141 Cervical vertebra_02 | 7.1 |  | 3.3 | 3.9 | 2.9 | 0.4 | 0.4 |
| 142 Cervical vertebra_03 | 40.7 | 13.7 | 8.6 | 18.3 | 13.7 | 2.2 | 1.8 |
| 143 Cervical vertebra_04 | 62.5 | 40 |  | 22.5 | 16.9 | 2.8 | 2.3 |
| 144 Cervical vertebra_05 | 47.5 | 36.1 |  | 11.4 | 8.5 | 1.6 | 1.2 |
| 145 Cervical vertebra_06 | 39.5 | 35.6 |  | 4 | 3 |  | 0.4 |
| 146 Cervical vertebra_07 | 8.6 | 8.6 |  |  |  |  |  |
| **Total** | **206.8** | **134.1** | **12.2** | **60.6** | **45.5** | **7.8** | **6.1** |

※1 Total body tissue = Cortical bone + Trabecular bone + Soft tissues
※2 Red bone marrow
※3 Red bone marrow 1,191.6 g, Blood 281.2 g, $281.2 \text{ g} \times 4/7/1191.6 \text{ g} = 13.5\%$
The blank cells indicate that no data is available.

**Table 2.** Percentage of blood distributed in the sinusoids calculated from bone marrow vessel data of mice.

| Column labels for calculations | a | b | c | d | e |
| --- | --- | --- | --- | --- | --- |
| Each structure and the geometrical conditions set for the calculation. | Vessel segments (n) | Mean diameter ($\mu\text{m}$) | Cross-sectional area of blood vessels $((b/2)^2 \times 3.14)$ ($\mu\text{m}^2$) | Cross-sectional area of each blood vessels $(c \times a)$ ($\mu\text{m}^2$) | Percentage of total cross-sectional area % |
| Arterial vessel | 9 | 8 | 50.2 | 452.2 | 3.7 |
| Post-arterial capillaries | 5 | 7.8 | 47.8 | 238.8 | 2.0 |
| Intermediate capillaries | 6 | 11.2 | 98.5 | 590.8 | 4.9 |
| Sinusoidal capillaries | 31 | 21.1 | 349.5 | 10,834.2 | 89.4 |
| **Total** |  |  |  | **12,116.0** |  |

Based on the statement of Saladine et al. ^(15)^ that sinusoids are typically 30–40 µm wide, the radius of the sinusoids was assumed to be 20 µm, and the number of vessels was assumed to be 40,000.

The total number of lattices was set at 1,600, the internal dimension of the lattice was set at 600 µm based on the data of Parfitt et al. ^(16)^, and the external dimension of the lattice was set to 630 µm based on the weight of the trabecular bone (Table 3, Figure 1).

**Figure 1.**
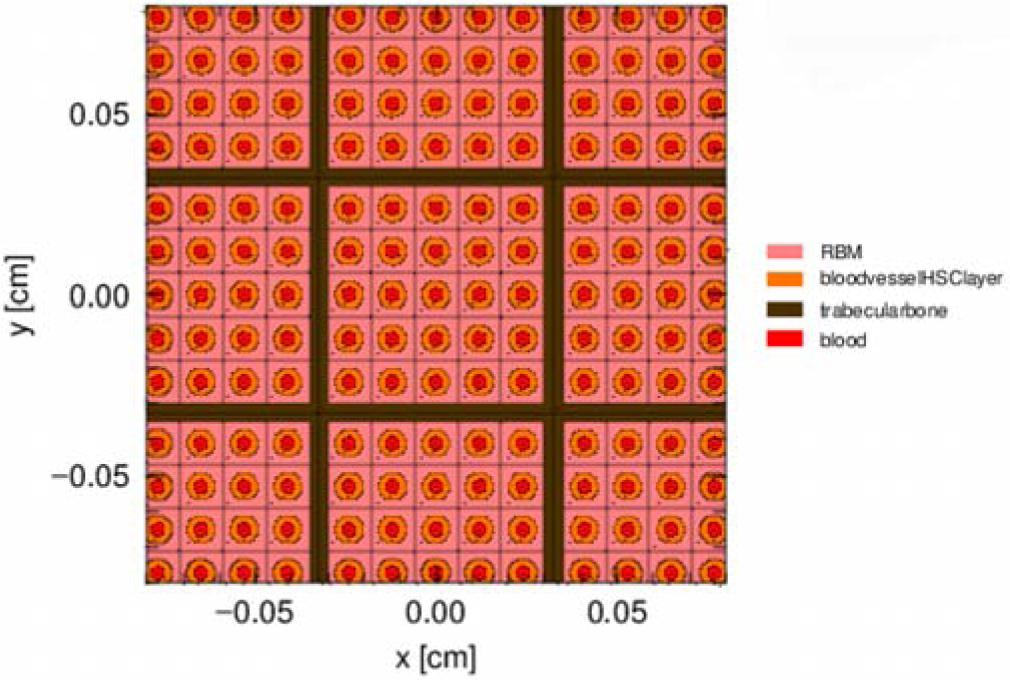
Geometry of the trabecular bone model constructed with the PHITS code

**Table 3.** Size of blood vessels and trabecular bone lattice calculated from the weight of bone tissue and blood volume.

| a | b | c | d | e | f | g | j | i |
| --- | --- | --- | --- | --- | --- | --- | --- | --- |
| RBM weight (g)<br>※1 | Blood weight (g)<br>※2 | Trabecular bone weight (g)<br>※3 | $b \times 89.4\%$<br>※4 | Number of sinusoids<br>※5 | Radius of sinusoids (cm)<br>※6 | Number of trabecular bone lattice<br>※7 | Size of RBM lattice (cm)<br>※8 | Size of Trabecular bone lattice (cm)<br>※9 |
| 45.5 | 6.1 | 12.2 | 5.5 | 40000 | 0.002 | 1600 | 0.06 | 0.063 |

- ※1 Weight of red bone marrow in cervical vertebra ※2 Weight of blood in cervical vertebra, calculated as 13.5% of RBM ※3 Weight of trabecular bone in cervical vertebra ※4 Weight of blood in sinusoids in cervical vertebra ※5 $=d / (0.002^2 \pi * 9)$ ※6 radius of sinusoids set based on data in Kenneth et al.<sup>(12)</sup> ※7 $=e/25$ ※8 $=\text{SQRT}(a/9/1600+f^2 \pi * 25)$ ※9 $=\text{SQRT}(j*j+c/1.765/1600/9)$

The target part of the organ was defined as the perivascular stem cell layer 10 µm from the vascular endothelium; Acar et al.^(7)^ reported that 85% of mouse HSCs were located within 10 µm of sinusoids, and Kunisaki et al.^(17)^ reported an average distance of 14.8 µm between the vascular endothelium and HSCs. The trabecular surface was defined as 10 µm from the surface of the trabecular bone and the inner 30 µm was defined as the trabecular volume. Because it is impossible to model the entire trabeculae, I modelled nine grids of 25 vessels each for a total of 225 vessels and multiplied the value obtained from the PHITS calculation by a factor of 40,000/225.

### 2.2 Radiation transport simulation and absorbed dose calculation

^137^Cs, ^131^I, ^90^Sr were selected for the calculation as beta-nuclides; ^223^Ra, ^239^Pu, ^238^U, ^232^Th, ^222^Rn as alpha-nuclides; and ^133^Xe, ^135^Xe, ^85^Kr as noble gases. Electron transport was simulated using the PHITS code ver. 3.17 for β-radionuclides and noble gases and alpha particles for alpha nuclides. The source regions were defined as blood, red bone marrow, trabecular bone volume, or trabecular bone based on the biokinetics of each radionuclide, and the target region was defined as the bone marrow stem cell layer, 10 µm from the vascular endothelium.

For each radionuclide, electrons or alpha particles were generated in the source region and the transferred energy distributed in the target region was calculated and converted to the absorbed dose per incident particle (Gy/source). For the calculation of beta nuclides, parameter type =28 was used for the source energy, which uses the DECDC ^(18)^ nuclear decay database (equivalent to ICRP107^(19)^) to obtain the energy spectra. The number of simulation trials was at least 10,000, and the statistical error in the target region was set to less than 0.05. For the alpha radionuclide, the statistical error was set to less than 0.5, owing to the long computation time required when using the trabecular bone as a source. For ^232^Th, the calculation was stopped with a statistical error of 0.9 because the energy distributed from the trabecular bone sources to the perivascular area was very small, which would have only a limited effect on the results and discussion, even though the statistical error is relatively large. The cut-off energy for the photons and electrons was set to 5keV. Bremsstrahlung was included in the simulation using EGS ^(20)^ mode.

### 2.3 Calculation of the number of decays in each compartment

Assuming that 1 Bq of radionuclide was inhaled, the number of decays in each compartment was calculated with R4.0.3^(21)^ using the deSolve code ^(22)^ and the transfer coefficients presented in ICRP 56^(23)^, 67^(24)^, and 69^(25)^ for the current model and those in ICRP 134^(26)^, 137^(27)^, and 141^(28)^ for the ICRP 103 model. The number of decays in each compartment of radionuclides transferred from the lungs to the blood was calculated for 15,800,000 min (10 years) for long-lived radionuclides and approximately 10,000 min for short-lived radionuclides. The choice of 10 years for long-lived nuclides instead of 50 years was made because of the limitations of the Personal Computer performance (Intel Core i5-3337U CPU 1.8 GHz, with 7.90 GB of RAM), and 10,000 minutes for short-lived radionuclides.

For noble gases, the ICRP presents only a kinetic model for radon dissolved in the blood vessels and transported into the body. Because xenon and krypton are relatively easy to distribute in fat^(29, 30)^ as is radon^(31)^, the transfer coefficients of radon were used for ^133^Xe, ^135^Xe, and ^85^Kr. Considering that the solubility of radon in water is twice that of xenon and four times that of krypton, it was assumed that 1/2 of xenon and 1/4 of krypton would be transferred to the blood.

The number of decays in red bone marrow blood was assumed to be proportional to the blood volume, which was 0.18% of the number of decays in whole-body blood (red bone marrow blood volume in the cervical spine (6.1 g)/ JM-103 blood (3,410 g) = 0.18%).

### 1.4 Calculation of the dose absorbed by the perivascular stem cell layer of the bone marrow after radionuclide inhalation

Assuming that 1 litre of air was inhaled after 1 h of exposure to air containing 1 Bq/m^3^ of radionuclides, the dose absorbed in the bone marrow perivascular stem cell layer was estimated by multiplying the absorbed dose determined in Section 2.2 by the decay number calculated in Section 2.3.

## 2. RESULTS

The absorbed doses calculated from the trabecular bone model and comparison with the specific absorbed fractions (SAFs) of ICRP60 and ICRP103 are shown in the tables.

### 3.1 Beta nuclides

The absorbed dose to the perivascular HSC layer from each source was calculated for ^137^Cs, ^131^I, and ^90^Sr, and compared with the doses estimated using the SAF and transfer coefficients in ICRP60 and ICRP103, presented in Tables 4-1, 4-2, and 4-3 as these are calculated using Microsoft Excel.

**Table 4-1.** Absorbed doses to the perivascular HSC layer for ^137^Cs calculated with the PHITS model and comparison with doses estimated using SAF and transfer coefficients in ICRP60 and ICRP103.

| $^{137}\text{Cs}$ - PHITS model | | | | | | | | |
| --- | --- | --- | --- | --- | --- | --- | --- | --- |
| a | b | c | d | e | f | g | h | i |
| source | Number of decay<br>Type M | Mass in cervical vertebrae (g) | Total mass in body (g) | Number of decay in cervical vertebrae | Absorbed dose to perivascular HSC layer for 225 vessels (Gy/source) | Absorbed dose to perivascular HSC layer for 40000 vessels (Gy/source) | Absorbed dose to perivascular HSC layer | Percentage of total absorbed dose % |
| Plasma | 54,326 | 6 | 3,410 | 778 | 8.84E-11 | 4.97E-13 | 3.87E-10 | 5% |
| Red blood cell | 381,394 |  |  |  |  |  |  |  |
| Red marrow | 407,957 | 45 | 1,192 | 15,401 | 7.72E-11 | 4.34E-13 | 6.69E-09 | 87% |
| TB surface | 406,381 | 12 | 2,466 | 1,978 | 5.32E-11 | 2.99E-13 | 5.92E-10 | 8% |
| Total |  |  |  |  |  |  | 7.67E-09 |  |

| $^{137}\text{Cs}$ - ICRP60 | | | | | |
| --- | --- | --- | --- | --- | --- |
| j | k | l | m | n | o |
| source | Number of decay<br>Type F | Absorbed dose per 1 MeV | Absorbed dose per 0.243 MeV | Absorbed dose (Gy/source) | Ratio of each organ (%) |
| Blood | 31,244 | 2.29E-15 | 5.56E-16 | 1.74E-11 | 0% |
| Body tissue A | 25,006 | 2.29E-15 | 5.56E-16 | 1.39E-11 | 0% |
| Body tissue B | 12,232,190 | 2.29E-15 | 5.56E-16 | 6.80E-09 | 100% |
| Total |  |  |  | 6.83E-09 | 100% |

| $^{137}\text{Cs}$ - ICRP103 | | | | | | |
| --- | --- | --- | --- | --- | --- | --- |
| p | q | r | s | t | u | v |
| Source | Number of decay<br>Type M | SAF to RBM for 0.2 MeV | Absorbed dose per 1MeV | Absorbed dose per 0.243MeV | Absorbed dose (Gy/source) | Percentage of total absorbed dose % |
| Plasma | 54,326 | 2.81E-02 | 4.50E-15 | 1.09E-15 | 5.94E-11 | 1% |
| Red blood cell | 381,394 |  |  |  | 4.17E-10 | 5% |
| Red marrow | 407,957 | 3.72E-01 | 5.95E-14 | 1.45E-14 | 5.90E-09 | 71% |
| TB surface | 406,381 | 1.29E-01 | 2.06E-14 | 5.00E-15 | 2.03E-09 | 23% |
| Total |  |  |  |  | 8.41E-09 | 100% |

- a: Source compartment b: Number of decays in each compartment based on the transfer coefficients of ICRP134 and ICRP137 c: Weight of tissue in the cervical vertebrae (g) d: Weight of tissue in the total body (g) e: Number of decays in the cervical vertebrae (b×c/d) f: Absorbed dose to the target per decay at the source calculated for 225 blood vessels. g: Absorbed dose per decay for the entire cervical vertebrae (f×225/40000) h: Absorbed dose to the target during the calculation period (g×e) i: Percentage of h to total absorbed doses in perivascular HSC layer j: Source compartment k: Number of decays in each compartment based on ICRP60's transfer coefficients l: Absorbed fraction per MeV (1 MeV×1.60 × 10<sup>-13</sup> (J MeV<sup>-1</sup>)×SAF, AF) m: Absorbed dose per decay (l×mean energy) n: Absorbed dose to the target during the calculation period (k×m) o: Percentage of n to total absorbed doses p: Source compartment q: Number of decays in each compartment based on ICRP103 recommendation r: SAFs provided in the SAF files of ICRP133 s: r× 1.6×10<sup>-13</sup> t: Absorbed dose per decay (s×mean energy) u: Absorbed dose to the target during the calculation period (q×t) v: Percentage of u to total absorbed doses

**Table 4-2.** Absorbed doses to the perivascular HSC layer for ^131^calculated with the PHITS model and comparison with doses estimated using SAF and transfer coefficients in ICRP60 and ICRP103.

| $^{131}\text{I}$ - PHITS model | | | | | | | | |
| --- | --- | --- | --- | --- | --- | --- | --- | --- |
| a | b | c | d | e | f | g | h | i |
| source | Number of decay<br>Type F | Mass in cervical vertebrae (g) | Total mass in body (g) | Number of decay in cervical vertebrae | Absorbed dose to perivascular HSC layer for 225 vessels (Gy/source) | Absorbed dose to perivascular HSC layer for 40000 vessels (Gy/source) | Absorbed dose in perivascular HSC layer | Percentage of total absorbed dose % |
| Blood 1 | 2,780 | 6 | 3,410 | 5 | 9.33E-11 | 5.25E-13 | 2.65E-12 | 62% |
| Blood 2 | 1,684 | 6 | 3,346 | 3 | 9.33E-11 | 5.25E-13 | 1.61E-12 | 38% |
| Total |  |  |  |  |  |  | 4.26E-12 | 100% |

| $^{131}\text{I}$ - ICRP60 | | | | | |
| --- | --- | --- | --- | --- | --- |
| j | k | l | m | n | o |
| source | Number of decay<br>Type F | Absorbed dose per 1 MeV | Absorbed dose per 0.1842 MeV | Absorbed dose (Gy/source) | Percentage of total absorbed dose % |
| Blood | 30,413 | 2.29E-15 | 4.21E-16 | 1.28E-11 | 100% |
| Total |  |  |  | 1.28E-11 | 100% |

| $^{131}\text{I}$ - ICRP103 | | | | | | |
| --- | --- | --- | --- | --- | --- | --- |
| p | q | r | s | t | u | v |
| Source | Number of decay<br>Type F | SAF to RBM for 0.2 MeV | Absorbed dose per 1MeV | Absorbed dose per 0.2MeV | Absorbed dose (Gy/source) | Percentage of total absorbed dose % |
| Blood 1 | 2,780 | 2.81E-02 | 4.50E-15 | 8.99E-16 | 2.50E-12 | 62% |
| Blood 2 | 1,684 | 2.81E-02 | 4.50E-15 | 8.99E-16 | 1.51E-12 | 38% |
| Total |  |  |  |  | 4.01E-12 | 100% |

**Table 4-3.**
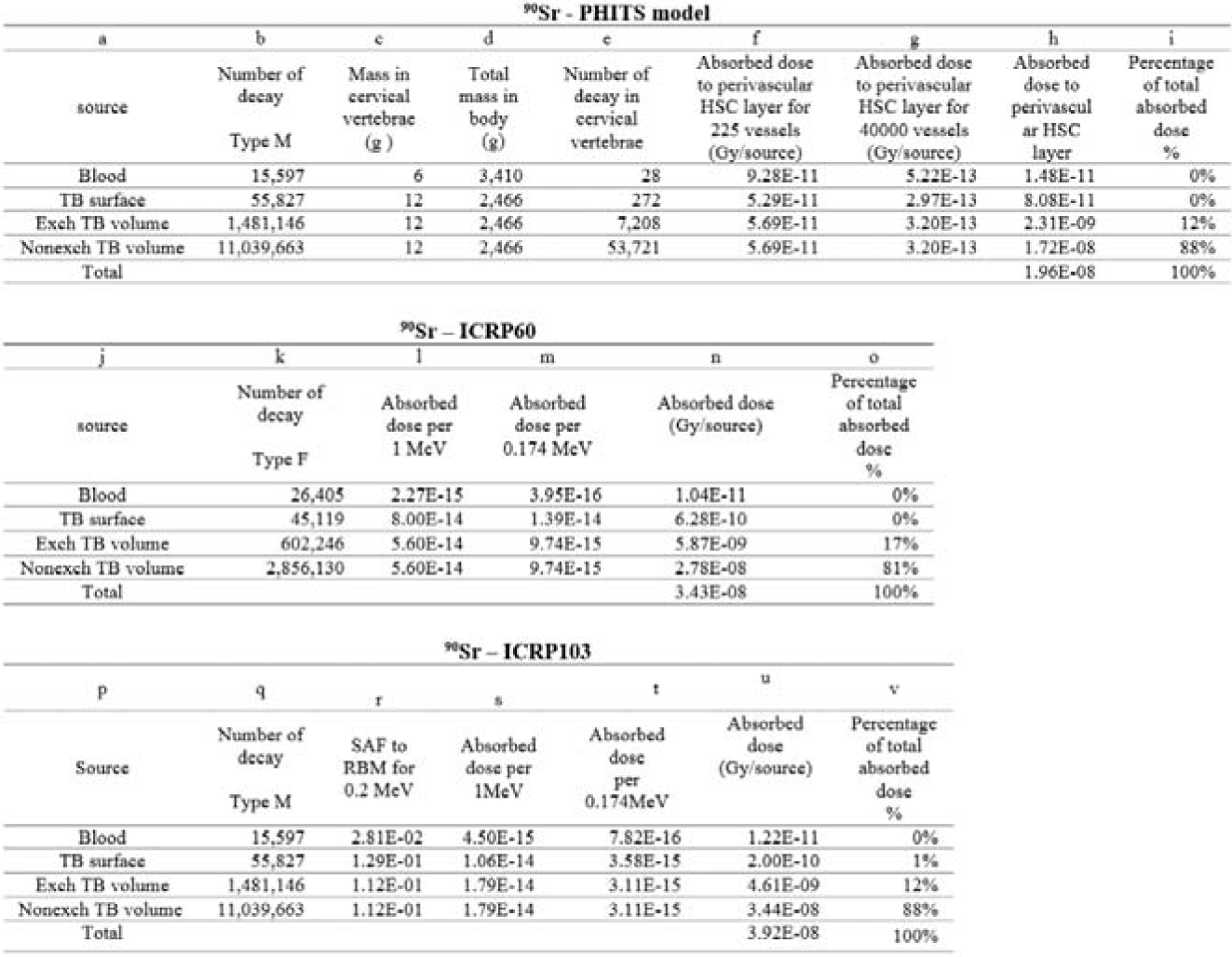
Absorbed doses to the perivascular HSC layer for ^90^Sr calculated with the PHITS model and comparison with doses estimated using SAF and transfer coefficients in ICRP60 and ICRP103.

### 3.2 Alpha nuclides

The calculation results for ^223^Ra, ^239^Pu, ^238^U, ^232^Th, ^222^Rn are presented in Tables 5-1, 5-2, 5-3, 5-4 and 5-5. As SAFs for radon are not provided in ICRP60 and ICRP103, only the results for the PHITS trabecular bone model are shown.

**Table 5-1.** Absorbed doses to the perivascular HSC layer for ^223^Ra calculated with the PHITS model and comparison with doses estimated using SAF and transfer coefficients in ICRP60 and ICRP103.

| <sup>223</sup>Ra - PHITS model |  |  |  |  |  |  |  |  |
| --- | --- | --- | --- | --- | --- | --- | --- | --- |
| a | b | c | d | e | f | g | h | i |
| Source | Number of decay<br>Type M | Mass in cervical vertebrae (g) | Total mass in body (g) | Number of decay in cervical vertebrae | Absorbed dose to perivascular HSC layer for 225 vessels (Gy/source) | Absorbed dose to perivascular HSC layer for 40000 vessels (Gy/source) | Absorbed dose to perivascular HSC layer | Percentage of total absorbed dose % |
| Blood | 2,270 | 6 | 3,410 | 4 | 7.49E-09 | 4.21E-11 | 1.74E-10 | 93% |
| TB surface | 5,442 | 12 | 2,466 | 26 | 8.53E-11 | 4.80E-13 | 1.27E-11 | 7% |
| Exch TB volume | 7,551 | 12 | 2,466 | 37 | 4.02E-12 | 2.26E-14 | 8.32E-13 | 0% |
| Nonexch TB volume | 568 | 12 | 2,466 | 3 | 4.02E-12 | 2.26E-14 | 6.25E-14 | 0% |
| Total |  |  |  |  |  |  | 1.88E-10 | 100% |

| <sup>223</sup>Ra – ICRP60 |  |  |  |  |  |
| --- | --- | --- | --- | --- | --- |
| j | k | l | m | n | o |
| source | Number of decay<br>Type M | Absorbed dose per 1 MeV | Absorbed dose per 5.979 MeV | Absorbed dose (Gy/source) | Percentage of total absorbed dose % |
| Blood | 538 | 2.27E-15 | 1.36E-14 | 7.31E-12 | 0% |
| TB surface | 7,176 | 8.00E-14 | 4.78E-13 | 3.43E-09 | 88% |
| Exch TB volume | 9,957 | 8.00E-15 | 4.78E-14 | 4.76E-10 | 12% |
| Nonexch TB volume | 748 | 8.00E-15 | 4.78E-14 | 3.58E-11 | 1% |
| Total |  |  |  | 3.92E-09 | 100% |

| <sup>223</sup>Ra – ICRP103 |  |  |  |  |  |  |
| --- | --- | --- | --- | --- | --- | --- |
| p | q | r | s | t | u | v |
| Source | Number of decay<br>Type M | SAF to RBM for 6 MeV | Absorbed dose per 1MeV | Absorbed dose per 5.979MeV | Absorbed dose (Gy/source) | Percentage of total absorbed dose |
| Blood | 2,270 | 2.87E-02 | 4.59E-15 | 2.74E-14 | 6.23E-11 | 8% |
| TB surface | 5,442 | 1.17E-01 | 1.87E-14 | 1.12E -13 | 6.10E-10 | 74% |
| Exch TB volume | 7,551 | 1.89E-02 | 3.02E-15 | 1.81E-14 | 1.36E-10 | 17% |
| Nonexch TB volume | 568 | 1.89E-02 | 3.02E-15 | 1.81E-14 | 1.02E-11 | 1% |
| Total |  |  |  |  | 8.19E-10 | 100% |

**Table 5-2.**
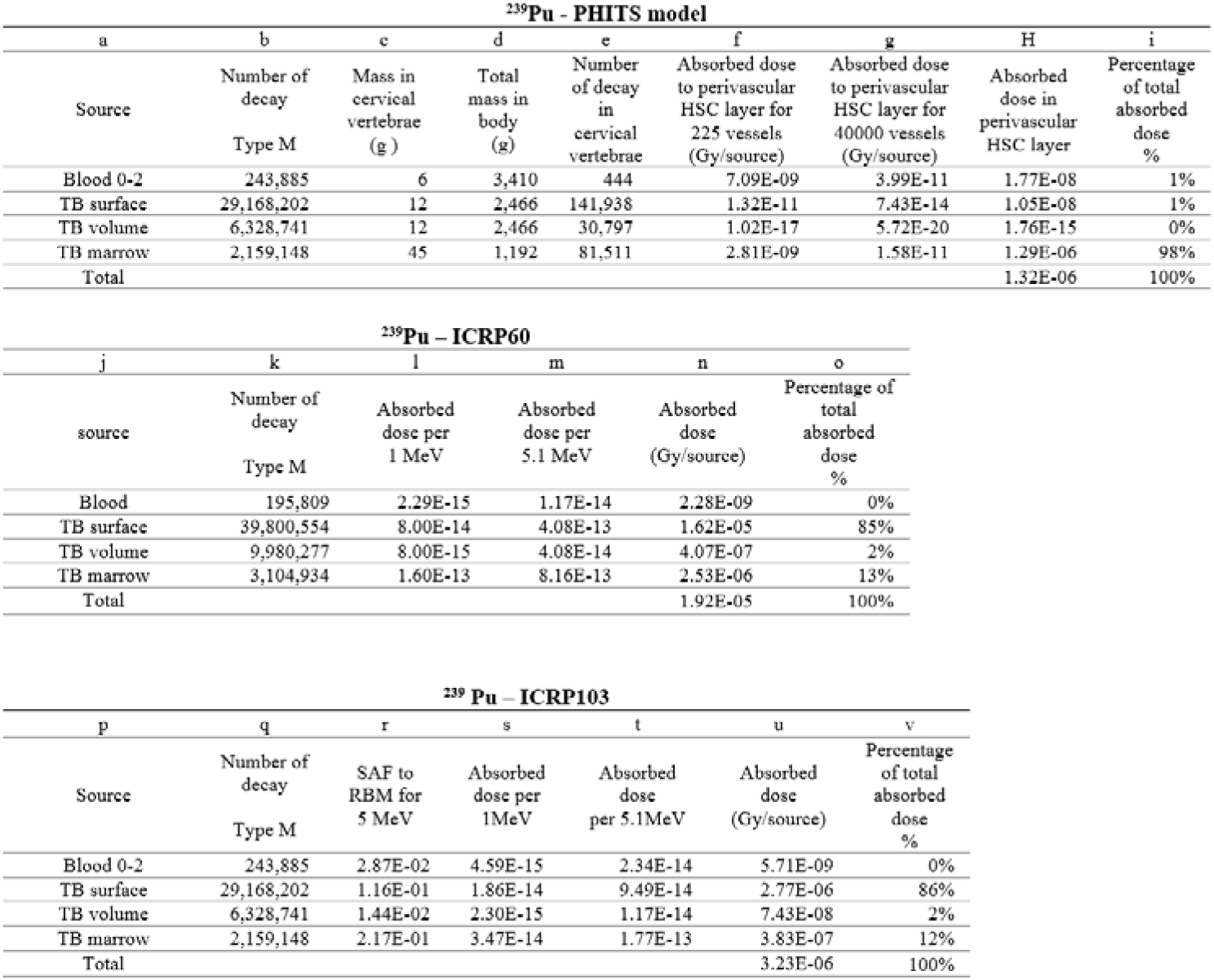
Absorbed doses to the perivascular HSC layer for ^239^Pu calculated with the PHITS model and comparison with doses estimated using SAF and transfer coefficients in ICRP60 and ICRP103.

**Table 5-3.**
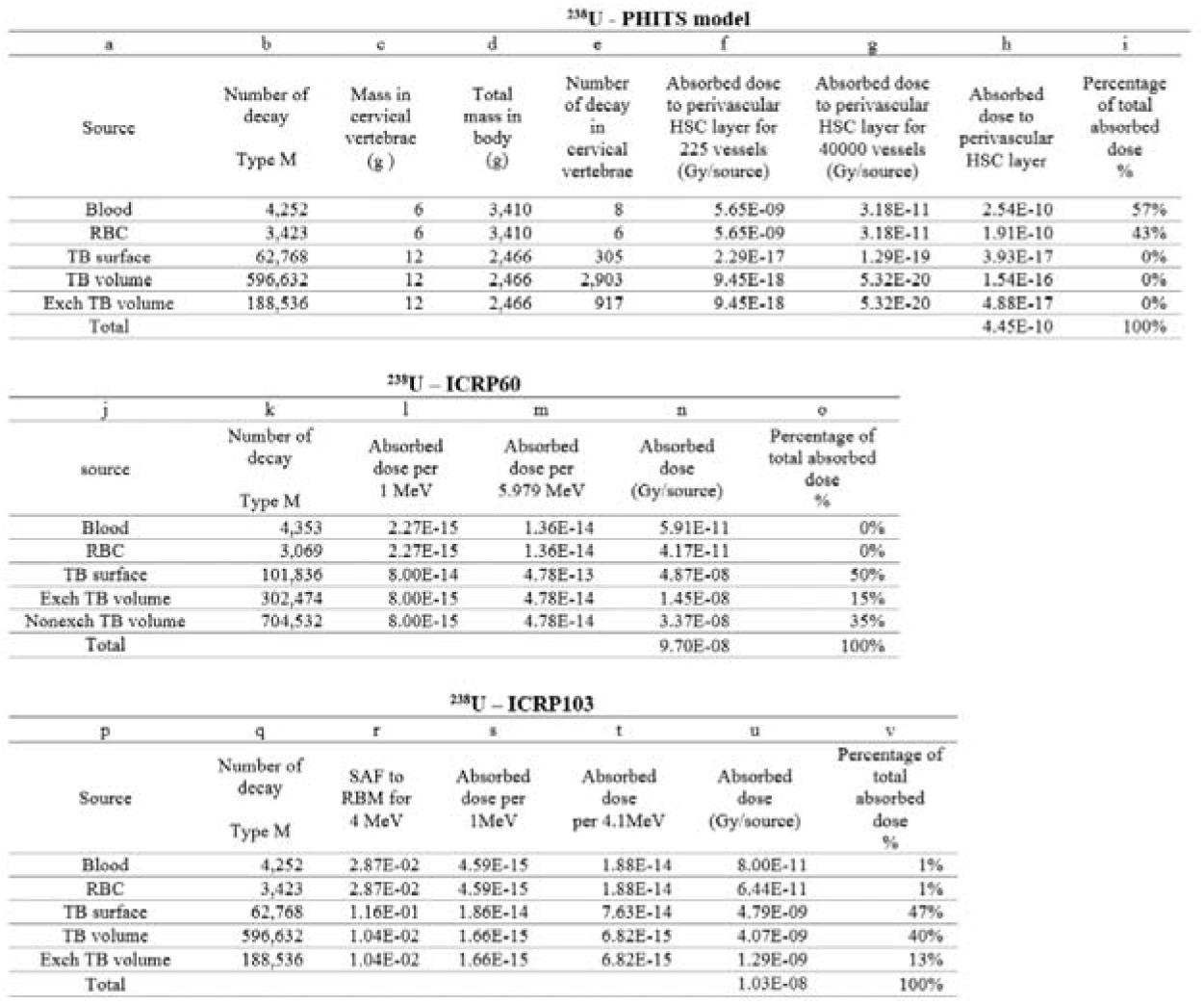
Absorbed doses to the perivascular HSC layer for ^238^U calculated with the PHITS model and comparison with doses estimated using SAF and transfer coefficients in ICRP60 and ICRP103.

**Table 5-4.** Absorbed doses to the perivascular HSC layer for ^232^Th calculated using the PHITS model and comparison with doses estimated using SAF and transfer coefficients in ICRP60 and ICRP103.

| <sup>232</sup>Th – PHITS model |  |  |  |  |  |  |  |  |
| --- | --- | --- | --- | --- | --- | --- | --- | --- |
| a | b | c | d | e | f | g | h | i |
| Source | Number of decay<br>Type S | Mass in cervical vertebrae (g) | Total mass in body (g) | Number of decay in cervical vertebrae | Absorbed dose to perivascular HSC layer for 225 vessels (Gy/source) | Absorbed dose to perivascular HSC layer for 40000 vessels (Gy/source) | Absorbed dose to perivascular HSC layer | Percentage of total absorbed dose % |
| Blood | 28,759 | 6 | 3,410 | 52 | 5.30E-09 | 2.98E-11 | 1.55E-09 | 0% |
| TB surface | 23,970,084 | 12 | 2,466 | 116,643 | 2.14E-19 | 1.20E-21 | 4.98E-16 | 0% |
| TB volume | 66 | 12 | 2,466 | 0 | 2.62E-19 | 1.48E-21 | 0 | 0% |
| Red marrow | 1,536,104 | 45 | 1,192 | 57,991 | 2.20E-09 | 1.10E-11 | 6.37E-07 | 100% |
| Total |  |  |  |  |  |  | 6.38E-07 | 100% |

| <sup>232</sup>Th – ICRP60 |  |  |  |  |  |
| --- | --- | --- | --- | --- | --- |
| j | k | l | m | n | o |
| source | Number of decay<br>Type M | Absorbed dose per 1 MeV | Absorbed dose per 4.1 MeV | Absorbed dose (Gy/source) | Percentage of total absorbed dose % |
| Blood | 78,306 | 1.60E-13 | 6.56E-13 | 5.14E-08 | 0% |
| TB surface | 57,703,246 | 8.00E-14 | 3.28E-13 | 1.89E-05 | 84% |
| TB volume | 16,519,053 | 8.00E-15 | 3.28E-14 | 5.42E-07 | 2% |
| TB marrow | 4,662,328 | 1.60E-13 | 6.56E-13 | 3.06E-06 | 14% |
| Total |  |  |  | 2.26E-05 | 100% |

| <sup>232</sup>Th – ICRP103 |  |  |  |  |  |  |
| --- | --- | --- | --- | --- | --- | --- |
| p | q | r | s | t | u | v |
| Source | Number of decay<br>Type S | SAF to RBM for 5 MeV | Absorbed dose per 1MeV | Absorbed dose per 5.1MeV | Absorbed dose (Gy/source) | Percentage of total absorbed dose % |
| Blood | 28,759 | 2.87E-02 | 4.59E-15 | 1.88E-14 | 5.41E-10 | 0% |
| TB surface | 23,970,084 | 1.16E-01 | 1.86E-14 | 7.61E-14 | 1.82E-06 | 79% |
| TB volume | 66 | 1.04E-02 | 1.66E-15 | 6.82E-15 | 4.50E-13 | 0% |
| Red marrow | 1,536,104 | 4.91E-01 | 7.86E-14 | 3.22E-13 | 4.95E-07 | 21% |
| Total |  |  |  |  | 2.32E-06 | 100% |

**Table 5-5.** Absorbed doses to the perivascular HSC layer for ^222^Rn calculated with the PHITS model.

| **<sup>222</sup>Rn - PHITS model** |  |  |  |  |  |  |  |  |
| --- | --- | --- | --- | --- | --- | --- | --- | --- |
| a | b | c | d | e | f | g | h | i |
| Source | Number of decay | Mass in cervical vertebrae (g) | Total mass in body (g) | Number of decay in cervical vertebrae | Absorbed dose to perivascular HSC layer for 225 vessels (Gy/source) | Absorbed dose to perivascular HSC layer for 40000 vessels (Gy/source) | Absorbed dose to perivascular HSC layer | Percentage of total absorbed dose % |
| Blood A | 4 | 6 | 3,410 | 0 | 7.40E-09 | 4.16E-11 | 0 | 0% |
| Blood V | 14 | 6 | 3,410 | 0 | 7.40E-09 | 4.16E-11 | 0 | 0% |
| TB volume | 1 | 12 | 2,466 | 0 | 4.09E-13 | 2.30E-15 | 0 | 0% |
| Red marrow | 31 | 45 | 1,192 | 1 | 3.01E-09 | 1.69E-11 | 1.69E-11 | 100% |
| Total |  |  |  |  |  |  | 1.69E-11 | 100% |

### 3.3 Noble gases

The results for ^133^Xe, ^135^Xe, and ^85^Kr are listed in Tables 6-1, 6-2 and 6-3. As the SAFs for noble gases are not provided in ICRP60 and ICRP103, only the results for the PHITS trabecular bone model are shown.

**Table 6-1.** Absorbed doses to the perivascular HSC layer for ^133^Xe calculated with the PHITS model.

| **<sup>133</sup>Xe - PHITS model** |  |  |  |  |  |  |  |  |
| --- | --- | --- | --- | --- | --- | --- | --- | --- |
| a | b | c | d | e | f | g | h | i |
| Source | Number of decay | Mass in cervical vertebrae (g) | Total mass in body (g) | Number of decay in cervical vertebrae | Absorbed dose to perivascular HSC layer for 225 vessels (Gy/source) | Absorbed dose to perivascular HSC layer for 40,000 vessels (Gy/source) | Absorbed dose to perivascular HSC layer | Percentage of total absorbed dose % |
| Blood A | 2 | 6 | 3,410 | 0 | 1.05E-10 | 5.99E-13 | 2.18E-15 | 1% |
| Blood V | 7 | 6 | 3,410 | 0 | 1.05E-10 | 5.99E-13 | 7.49E-15 | 3% |
| TB volume | 0 | 12 | 2,466 | 0 | 0 | 0 | 0 | 0% |
| Red marrow | 15 | 45 | 1,192 | 1 | 7.15E-11 | 4.02E-13 | 2.28E-13 | 96% |
| Total |  |  |  |  |  |  | 2.37E-13 | 100% |

**Table 6-2.**
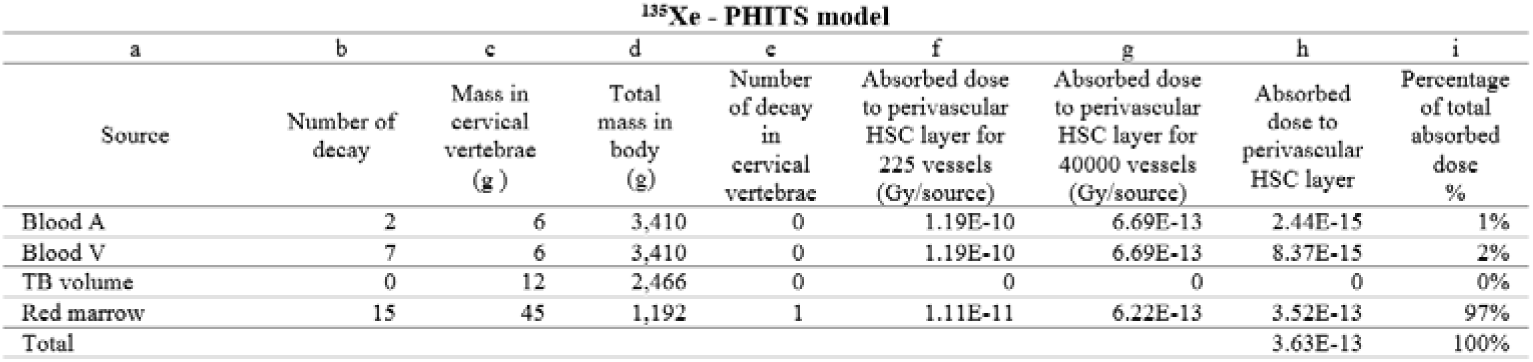
Absorbed doses to the perivascular HSC layer for ^135^Xe calculated with the PHITS model.

**Table 6-3.** Absorbed doses to the perivascular HSC layer for ^85^Kr calculated with the PHITS model.

| <sup>85</sup>Kr - PHITS model |  |  |  |  |  |  |  |  |
| --- | --- | --- | --- | --- | --- | --- | --- | --- |
| a | b | c | d | e | f | g | h | i |
| Source | Number of decay | Mass in cervical vertebrae (g) | Total mass in body (g) | Number of decay in cervical vertebrae | Absorbed dose to perivascular HSC layer for 225 vessels (Gy/source) | Absorbed dose to perivascular HSC layer for 40,000 vessels (Gy/source) | Absorbed dose to perivascular HSC layer | Percentage of total absorbed dose % |
| Blood A | 1 | 6 | 3,410 | 0.0 | 1.04E-10 | 5.87E-13 | 1.07E-15 | 1% |
| Blood V | 4 | 6 | 3,410 | 0.0 | 1.04E-10 | 5.87E-13 | 4.19E-15 | 3% |
| TB volume | 0 | 12 | 2,466 | 0 | 0 | 0 | 0 | 0% |
| Red marrow | 8 | 45 | 1,192 | 0.3 | 9.41E-11 | 5.29E-13 | 1.60E-13 | 97% |
| Total |  |  |  |  |  |  | 1.65E-13 | 100% |

## 3. DISCUSSION

Table 7 summarizes the total absorbed doses to the perivascular HSC layer obtained from the PHITS calculation for each nuclide and a comparison with the ICRP60 and ICRP103 estimates. For β-nuclides, the absorbed dose per decay was higher in the PHITS model for all three nuclides, but the absorbed dose was almost the same as that in ICRP60 because the number of decays in each compartment changed significantly owing to changes in the biokinetic model and transfer coefficients.

**Table 7.** Summary of the calculated absorbed doses to the perivascular HSC layer.

|  | nuclides | PHITS model<br>(Gy/source) | ICRP60<br>(Gy/source) | ICRP103<br>(Gy/source) | ICRP60/PHITS | ICRP103/PHITS |
| --- | --- | --- | --- | --- | --- | --- |
| beta-nuclides | $^{137}\text{Cs}$ | 7.67E-09 | 6.83E-09 | 8.41E-09 | 0.9 | 1.1 |
| | $^{131}\text{I}$ | 4.26E-12 | 1.28E-11 | 4.01E-12 | 1.8 | 0.9 |
| | $^{90}\text{Sr}$ | 1.96E-08 | 3.43E-08 | 3.92E-08 | 1.4 | 2.0 |
| alpha-nuclides | $^{223}\text{Ra}$ | 1.88E-10 | 3.92E-09 | 8.19E-10 | 20.9 | 4.4 |
| | $^{239}\text{Pu}$ | 1.32E-06 | 1.92E-05 | 3.23E-06 | 14.6 | 2.5 |
| | $^{238}\text{U}$ | 4.45E-10 | 9.70E-08 | 1.03E-08 | 217.8 | 23.1 |
| | $^{232}\text{Th}$ | 6.38E-07 | 2.26E-05 | 2.32E-06 | 35.4 | 3.6 |
| | $^{222}\text{Rn}$ | 1.69E-11 | - | - | - | - |
| noble gases | $^{133}\text{Xe}$ | 2.37E-13 | - | - | - | - |
| | $^{135}\text{Xe}$ | 3.63E-13 | - | - | - | - |
| | $^{85}\text{Kr}$ | 1.65E-13 | - | - | - | - |

For alpha nuclides, few particles reached the perivascular HSC layer from the trabecular bone (TB) source because of their short range (Figure 2, nuclide: ^239^Pu, source organ: TB surface). This is consistent with a report by Tranel et al. that the range of alphas ensures that the absorbed dose is minimal at distances greater than 100μm^(10)^. Most of the dose to the perivascular HSC layer came from either the red bone marrow source or the blood source; therefore, the dose calculated by the PHITS model was lower than the dose assessment based on the ICRP 60 recommendation, which assumes an AF of 0.5 for the source TB surface and 0.05 for the TB volume. Compared to the dose estimated using the ICRP133 SAF, the difference was smaller, approximately 2–23 times lower. An evaluation of alpha nuclides using a more accurate model is required.

**Figure 2.**
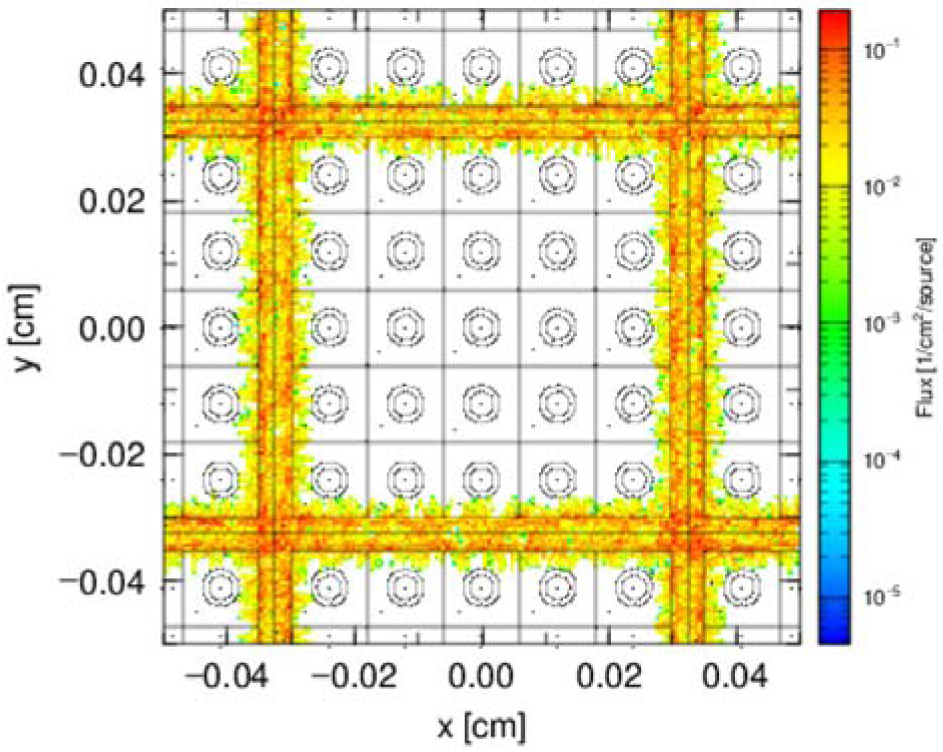
PHITS simulation of ^239^Pu deposited on the trabecular bone surface to the perivascular HSC layer

If HSCs are located in the perivascular HSC layer of the red bone marrow and are less susceptible to alpha radionuclides in bone sources, as suggested in Tables 5-1, 5-2, 5-3 and 5-4, the internal doses of alpha nuclides in epidemiological studies to date have been overestimated, and the actual doses to the red bone marrow may be lower. ^223^Ra has been used for the treatment of prostate cancer, and red bone marrow doses have been evaluated by Lassmann et al.^(32)^, the actual doses may be lower. Similar to ^222^Rn, red bone marrow sources affect the stem cell layer. Radon biokinetics and bone marrow absorbed doses must be evaluated, as reported by Sakota et al.^(33)^.

Noble gases, for which internal exposure is not currently assessed, have lower absorbed doses per decay than beta and alpha nuclides; however, exposure to large quantities may have radiation effects on the bone marrow stem cell layer. This may contribute to the radiation exposure of people living near accidents and nuclear power plants, where the effects of radiation exposure are controversial. A large amount of ^133^Xe was released into the environment during the Three Mile Island accident in 1979, but the exposure of nearby residents to xenon was assessed only for external exposure. Internal exposure was not included in the radiation doses. Datesman ^(34)^ noted a discrepancy between the results of physical dosimetry and biodosimetry by cytogenetic analysis of residents living near the Three Mile Island NPP. Noble gases are 10 times more soluble in lipids than in non-lipid tissues^(35)^, and Wang et al.^(36)^ reported that bone marrow fat accounts for approximately 10% of the total fat in healthy adults. It has also been reported that bone marrow adipocytes are located adjacent to sinusoidal blood vessels and are hematopoietic^(37)^. The biokinetics of xenon and other noble gases in the body and the assessment of their exposure to the bone marrow stem cell layer should be considered.

In terms of limitations, the trabecular bone model used in this study is a simple model of part of the cervical vertebrae, although it is based on available human data. A precise model based on micro-computed tomography (CT) images is required for dosimetry. Additionally, because the transfer coefficients for noble gases are estimated from the coefficients for radon, it is necessary to construct a pharmacokinetic model based on actual measurements.

## 4. CONCLUSION

The bone marrow doses calculated using the PHITS trabecular bone marrow model, which assumes that the stem cell layer is located in the perivascular HSC layer of the sinusoids, showed that the absorbed doses from the bone marrow and blood sources were greater than those from trabecular bone sources for alpha nuclides, and the total absorbed dose was lower than that estimated from the current ICRP models. Bone marrow dose assessments from internal exposure should be re-examined using a more detailed model of the trabecular bone marrow cavity, assuming a heterogeneous distribution of HSCs and other bone marrow cells. It is also necessary to assess the effects of fat-soluble noble gases on HSCs in the bone marrow from the perspective of occupational and public radiation protection.

## Data Availability

All data produced are available online at

https://docs.google.com/spreadsheets/d/1nfSBkNYn8jN18dX5mqZNJrcgrWiNTxlveK2K-zKQiWs/edit?gid=0#gid=0

## ACKNOWLEDGEMENTS

This paper was written by a Japanese citizen who learned the ICRP’s internal radiation exposure assessment methodology after the Fukushima nuclear accident with the aim of ending the controversy over radiation exposure. I would like to express my gratitude and deepest respect to Ichiro Yamaguchi, National Institute of Public Health, Japan, for his support and efforts in risk communication for over 10 years. He answered my questions and supported the writing of this paper; however, all assertions, conclusions, and mistakes herein are the sole responsibility of the author. Sincere thanks are also due to Seiko Hirota and other members of the Young Researchers Association of JHPS for allowing me to join the study group and to the reviewer of JHPS who provided me with accurate and valuable comments. I would also like to thank Takumi Goto and Hidenaga Yoshioka for their support and assistance via the internet. I hope that experts will address the issues raised in this study. This study is not funded by any institutions and the author declare no conflicts of interest.

